# Copy Number Variant analysis by exome sequencing is an effective approach to optimize diagnostic yield for developmental disorders – the DDD-Africa study

**DOI:** 10.64898/2026.02.06.26345639

**Authors:** Nadja Louw, Prince Makay, Phelelani.T Mpangase, Thirona Naicker, Laura M. Yates, Engela Honey, Gerrye Mbungu, Kris Van Den Bogaert, Helen V. Firth, Matthew E. Hurles, Prosper Lukusa Tshilobo, Koen Devriendt, Amanda Krause, Nadia Carstens, Aimé Lumaka, Zané Lombard

## Abstract

Copy number variants (CNV) contribute significantly to the pathogenic variation associated with developmental disorders. CNV detection is often not included in standard exome sequencing (ES) analysis. Complementary methods such as chromosomal microarray are typically offered in diagnostic laboratories to diagnose pathogenic CNV. In this study, we aimed to develop an optimal approach for incorporating CNV detection within our ES analysis process for the Deciphering Developmental Disorders in Africa (DDD-Africa) cohort. We analyzed ES data from 505 probands with a developmental disorder, applying a CNV detection approach that assessed data generated using the tools CANOES and XHMM. When available, parental ES data was used to assess inheritance patterns. We confirmed a diagnosis in 42/505 (8,3%) patients with 44 pathogenic CNV identified in the probands. There were 31 deletions and 13 duplications. Among the 27 probands with parental data, all identified CNV were *de novo*. The addition of CNV analysis to our ES analysis pipeline resulted in an 8.3% increase in diagnostic yield in the DDD-Africa cohort without additional laboratory cost. This approach offers a feasible approach which is likely to reduce analytical cost and is suitable for low- and middle-income countries where funding and resources for genomic medicine initiatives are limited.

## Introduction

Developmental disorders (DD) are challenging to diagnose due to both clinical and genetic heterogeneity. This is even more challenging in low-middle income countries (LMICs) where fewer clinical and laboratory resources are available for genomic medicine interventions. Exome sequencing (ES) has proven to be a cost-effective first-tier test for genetic diagnosis of DD in developed countries, predicting significant cost saving per diagnosis (1). As a result, ES is also an attractive option for diagnostic implementation in LMICs, although the lack of established infrastructure and trained personnel remains a consideration. In addition, the ability to detect both single nucleotide variation (SNV) and copy number variation (CNV) using a single testing approach further enhances its potential utility in a limited resourced environment.

Copy number variations are defined as deletions and duplications with a size greater than 50bp (2, 3). Large pathogenic CNV (>100kb) are causal in at least ∼15% of DD patients (4–6). Chromosomal microarray (CMA) is still widely used diagnostically for CNV detection but requires specialized equipment. The approach of combining SNV and CNV detection from ES data is potentially more cost-effective as only one technology is used. The implementation of this integrated approach has been shown to work well in resource limited settings although it has only been studied in a limited number of LMICs where it increased the diagnostic yield by an average of 10.6% compared to SNV-only ES detection (7–14).

There is currently no gold standard detection method for CNV from ES data. Analysis relies mainly on depth of coverage comparison data, but no single approach calls CNV with high sensitivity and specificity (15). Therefore, an active area of research for optimized CNV detection from NGS data is to use multiple tools in parallel and then combine the data (16). It could therefore be advantageous to investigate and establish an approach for CNV detection from ES data, particularly in LMICs.

In this study, we applied two different well-established bioinformatics CNV calling tools with different algorithmic approaches to detect CNV from ES data on the Deciphering Developmental Disorders in Africa study (DDD-Africa) dataset. The aim of DDD-Africa is the implementation of genomic medicine solutions for rare developmental disorders in Africa, with a focus on cohorts from South Africa (SA) and the Democratic Republic of Congo (DRC). We assess the added diagnostic yield of ES-based CNV calling and its utility within a resource limited environment.

## Materials and Methods

### Participants

The DDD-Africa study was initiated with one of its aims being to evaluate the implementation of ES as a first-tier genetic diagnostic test for DD in Africa (17). Ethics approval was obtained for this study through the Human Research Ethics Committee - Medical of The University of The Witwatersrand (certificate number: M230567), University of Pretoria (80/2018) and the University of Kwazulu-Natal (RECIP006/2020) within South Africa as well as from the University of Kinshasa (ESP/CE/050/2018) in the DRC.

The DDD-Africa cohort consists of 505 probands with DD who had no prior genetic diagnosis, and their parents (where available). The average age of the probands was 7.2 years (SD=5.3), with 62% (315/505) male and 38% (190/505) female. In 17 cases, more than one similarly affected individual was recruited as an extended family. Families were recruited from study sites in either South Africa (358 probands) or the DRC (147 probands). A total of 477 mothers and 259 fathers were enrolled in the study, resulting in a final dataset of 239 trios, 235 duos, 17 extended families with more than one affected individual and 14 singletons (N=1263 participants). A total of 315/505 probands were male with 97/147 recruited from DRC and 218/358 recruited from SA. The remaining probands (190/505) were female with 50/147 recruited form DRC and 140/358 recruited from SA. Detailed clinical phenotype information and DNA samples were collected followed by ES analysis. The majority of SA participants (94.6%) had some genetic testing prior to recruitment, compared to only 8.2% of participants from the DRC. In the South African cohort, this included karyotype analysis and MLPA for common microdeletion/duplication syndromes and subtelomeric deletions and duplications. Chromosomal microarray analysis was also completed for a small subset.

The participant inclusion criteria were adapted from the DDD-UK study which aimed to include children with severe undiagnosed DD (18). All individuals matching at least one of the inclusion categories below were eligible for enrolment into the study:

I. Individuals with moderate to severe developmental delay of unknown cause.
II. Individuals with mild developmental delay, and additional clinically relevant minor anomalies or dysmorphic features.
III. Individuals with major malformations in two or more different organ systems.
IV. Individuals with major malformation in one organ system and additional clinically relevant minor anomalies or dysmorphic features.

### Exome Sequencing

ES was performed at the Wellcome Sanger Institute (Hinxton, UK) using ISC-Twist library preparation (Twist Biosciences, San Francisco, CA, USA). Samples were processed in pools of 96-plex and sequenced using Illumina paired-end technology (Illumina, San Diego, CA, USA) on the Illumina NovaSeq 6000 platform, targeting an average depth of coverage of ∼40x. Although this coverage is lower than the typical coverage of traditional diagnostic ES approaches (∼100x), it has been shown to yield sufficiently comparable variant detection ability (19). This approach has also been successfully implemented in the large-scale DDD-UK study (18).

### Bioinformatic CNV calling from ES data

In order to try and optimize the bioinformatic CNV tool or combination of tools, a separate validation study (20) was completed prior to using the CNV tools on all DDD-Africa participants. In the validation study, using a subset of DDD-Africa samples, three freely accessible CNV calling tools, namely CANOES (21), CLAMMS (22) and XHMM (23), were assessed to identify the best tool or combination for implementation on the ES data. These tools were chosen as they use different statistical models for CNV detection and were also used as part of the CNV analysis for the DDD-UK study (24). The validation study revealed that the combined use of CANOES and XHMM provided optimal detection of likely disease-causing CNV. CANOES uses a negative binomial distribution to call CNV from autosomes only, which has been shown to be a good model for overdispersed sequence depth data and is focused on calling rare CNV of 100kb-10Mb. XHMM uses a Gaussian approach to call CNV and is specifically designed for, but not limited to, large cohorts. XHMM detects novel, rare CNV (<5% of cohort) over 200kb in size and can detect CNV on both autosomes and sex chromosomes. CLAMMS did not significantly increase the CNV detection rate and was therefore not applied to the main dataset.

A Nextflow pipeline was created and tools were containerized using Docker (25). All CNV calling tools were applied using default settings.

### CNV classification and filtering

CNV were classified using the American College of Medical Genetics and Genomics (ACMG) and the Clinical Genome Resource (ClinGen) technical standards (26). This was done using ClassifyCNV (27) from the web-based application CNV-ClinViewer (28). CNV detected by each tool were uploaded separately into CNV-ClinViewer. Following classification, filtering was applied based solely on the proband genomic data. CNV observed in parental data were only used to establish inheritance patterns of shortlisted CNV. Specific filtering steps were needed to ensure technical validity of the CNV. Firstly, to minimize false positive results, classified CNV were filtered according to each tool’s quality metrics, specifically the phred-scaled quality score indicating the probability of a CNV being present within the interval (Q_SOME) for CANOES (>80), and XHMM (>60) (Figure 1). CNV were also filtered by size (>100kb). The size cutoff was introduced given that CNV larger than 100kb in size are more likely to be pathogenic (5, 29). This cutoff was used as a prioritization step rather than an exclusion criterion as smaller CNV that were classified as likely pathogenic and pathogenic (LP/P) were also investigated. After classification and filtering, all LP/P CNV were categorized by size into different groups (>1Mb, 500kb-1Mb and 100-500kb). The filtered LP/P variants for each tool were shortlisted and compared to obtain a final list of plausible disease-associated CNV. LP/P CNV identified by both tools were prioritized before analyzing LP/P CNV called with only one tool. A genotype-phenotype correlation was carried out by clinical geneticists on final shortlisted variants with results confirmed at a multi-disciplinary variant review meeting. DECIPHER (30) was used by the DDD-Africa project to share and compare phenotypic and genotypic data, and therefore all shortlisted CNV were uploaded to this database. Participants are referred to by their DECIPHER ID in this manuscript for ease of reference.

**Figure 1:**
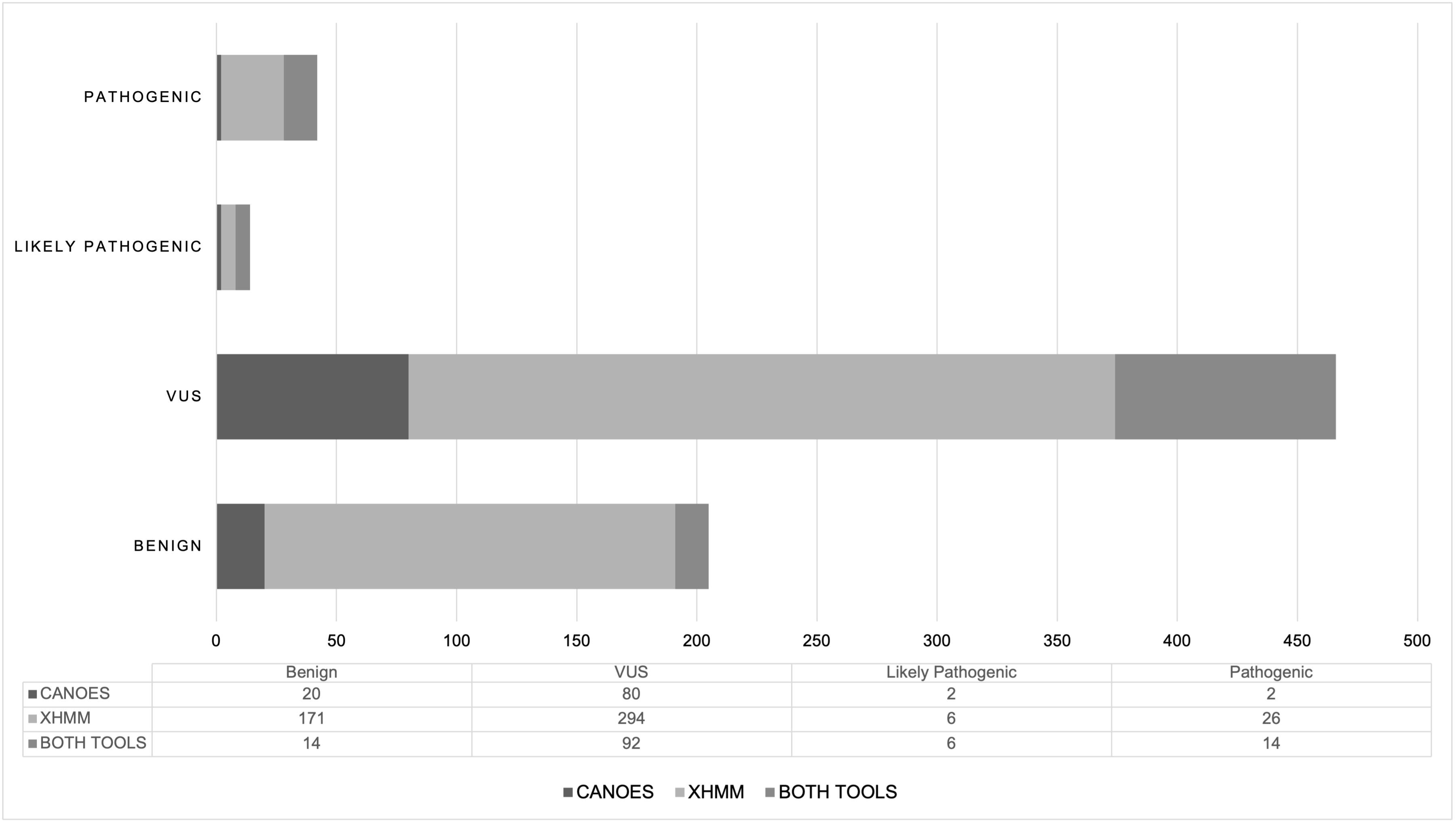
Web-based classification of CNVs. Manual filtering (step 2) was carried out in three steps included selecting probands only, larger CNVs (>100kb) and applying specific quality scores for each tool (2A, 2B and 2C).

### CNV validation using Array CGH

In total, CNV were validated by array CGH for 19/42 participants. Eleven of the 19 CNV were from the SA cohort and were confirmed using the Agilent SurePrint G3 ISCA v2 CGH 8×60K array (Agilent Technologies, Santa Clara, CA, USA) as per manufacturer’s instructions. Agilent CytoGenomics 5.3 was used for data analysis with reference tracks from DGV, International Standards for Cytogenomic Arrays, OMIM and in-house data. Array analysis was performed as confirmation prior to returning patient results. The array CGH was performed in a diagnostic laboratory by the Division of Human Genetics, National Health Laboratory Service (NHLS), Johannesburg, South Africa.

Eight of the 19 validated CNV were from the DRC cohort and were confirmed using the 4×180K CytoSure ISCA v3 array (Oxford Gene Technology, Oxford, Oxfordshire, UK) as per manufacturer’s instructions. Data visualization and analysis were performed with CytoSure Interpret Software (OGT) using the circular binary segmentation algorithm. The DRC samples were selected where the exact nature of the CNV could not unambiguously be defined without independent validation. These selected CNV included complex rearrangements (multiple CNV in one individual), suspected trisomy of an entire chromosome or chromosome arm or CNV for which verification of involvement of a critical gene was required. Array analysis was performed at the Laboratory for Cytogenetics and Genome Research, Center for Human Genetics, KU Leuven, Belgium.

### Statistical analysis

The MedCalc software was used to calculate p-values a Chi-squared test for the comparison of two proportions (from independent samples), expressed as a percentage. [MedCalc Software Ltd. Comparison of proportions calculator. https://www.medcalc.org/calc/comparison_of_proportions.php (Version 23.3.7; accessed August 25, 2025)].

## Results

A total of 44 pathogenic CNV was identified from 42/505 (8.3%) probands after ES CNV analysis. Most of the probands with a LP/P CNV detected were female (25/42). The diagnostic yield showed a significant difference (p<0.05) between the SA cohort at 6.1% (22/358) and the DRC cohort at 13.6% (20/147).

Raw CNV detection data yielded 61166 CNV (Figure 2). After filtering for CNV present in probands only, 23059 CNV were shared between the two tools. A total of 6282 CNV were detected from probands by CANOES before any quality or size filtering was implemented. The majority were deletions (70.8%) with an average size of 1.5Mb. The average size of duplications (29.2%) was 0.1Mb. More than double the number of CNV were detected by XHMM from probands of which 52.1% were duplications with an average size of 0.1Mb, whereas the deletions (47.9%) had an average size of 0.09Mb.

**Figure 2:**
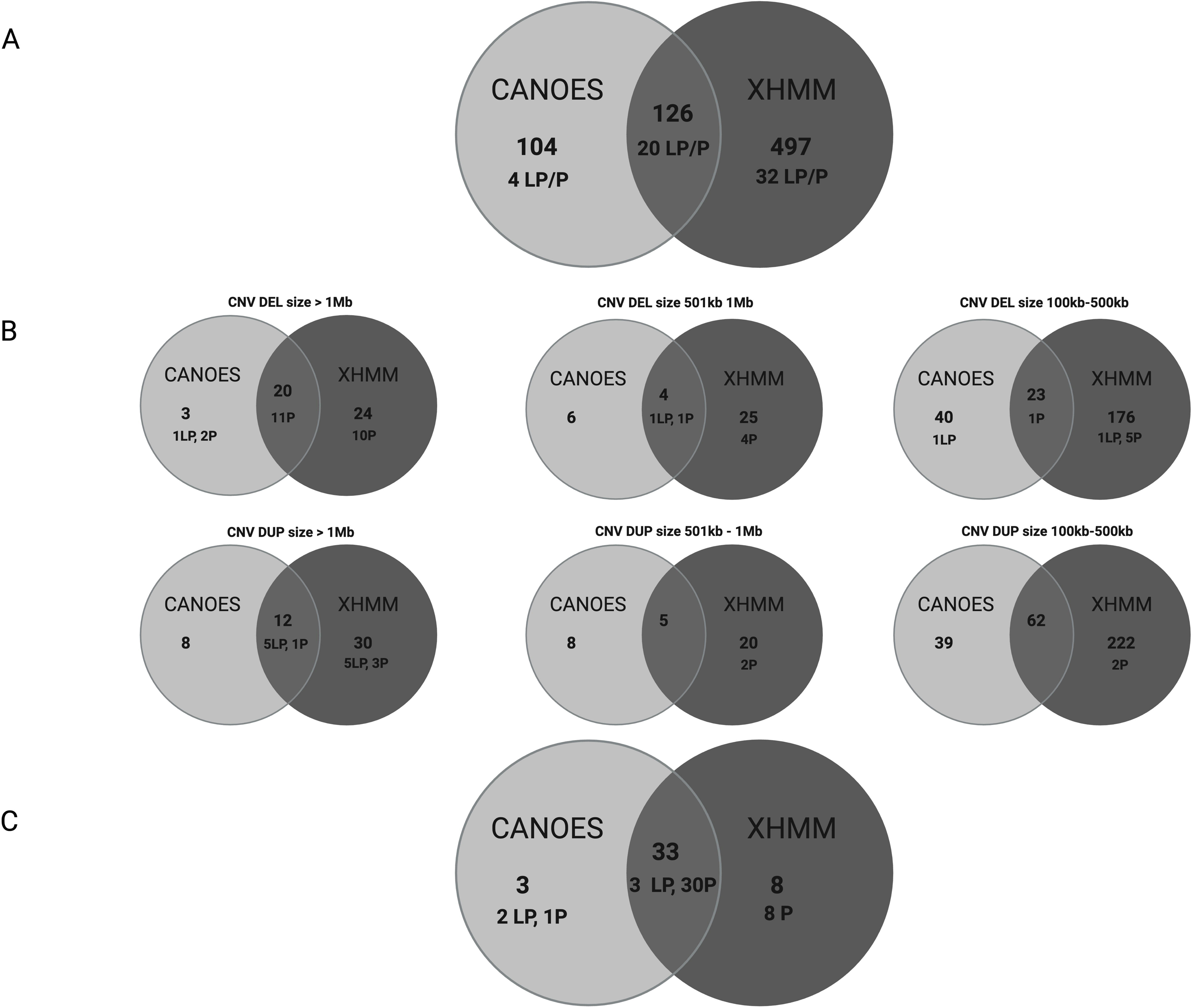
Filtering of CNVs detected by CANOES and XHMM. Starting from raw data, CNVs only present in probands were selected followed by CNVs meeting quality criteria (phred scores) and lastly filtering out CNVs with identical intervals present in multiple probands and unaffected individuals and/or population databases.

After filtering to ensure CNV met the quality parameters and to select for CNV >100kb, a total of 3474 CNV were retained. Additional filtering was performed to exclude all CNV with identical breakpoints and size present in more than two individuals as well as in population databases thus not likely to be disease-causing. CNV present in >2 individuals with identical breakpoints in regions of interest and not present in the population databases as part of normal variation were not excluded.

This led to the inclusion of 727 CNV (Figure 2) of which the majority were classified as Variants of Uncertain Significance (VUS), 62% of XHMM CNV and 74% of CNV from CANOES (Figure 3). A total of 56 CNV were classified as likely pathogenic or pathogenic (LP/P) across both tools, of which 20 were identified by both CANOES and XHMM.

**Figure 3:**
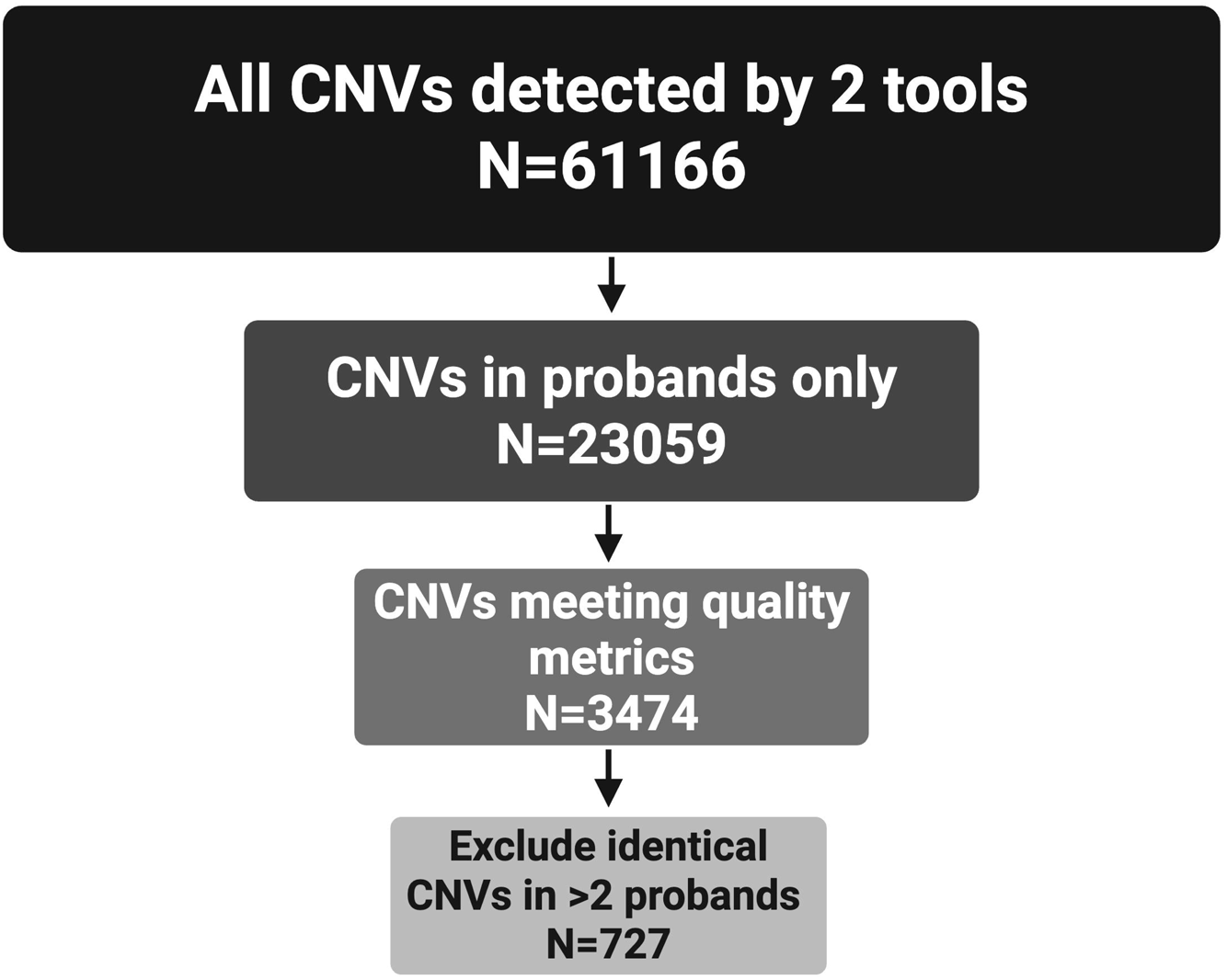
Disease impact categories for the unique CNVs identified in probands using CANOES (charcoal), XHMM (light grey) and both tools (dark grey). (VUS = Variant of uncertain Significance)

A total of 126 CNV were detected by both tools (Figure 4A). Further filtering of these CNV by size (Figure 4B) showed that over 50% of LP/P CNV are deletions >1Mb. Focusing on CNV below 100kb (not included in Figure 4), ten CNV classified as LP/P by CNV-ClinViewer were detected by XHMM of which only six met the quality criteria. Only one of the six was shortlisted as a LP/P CNV after genotype-phenotype correlation was completed. CANOES detected 27 LP/P CNV <100kb of which only three met quality criteria resulting in one LP CNV included in the final shortlist (Figure 4C).

**Figure 4.**
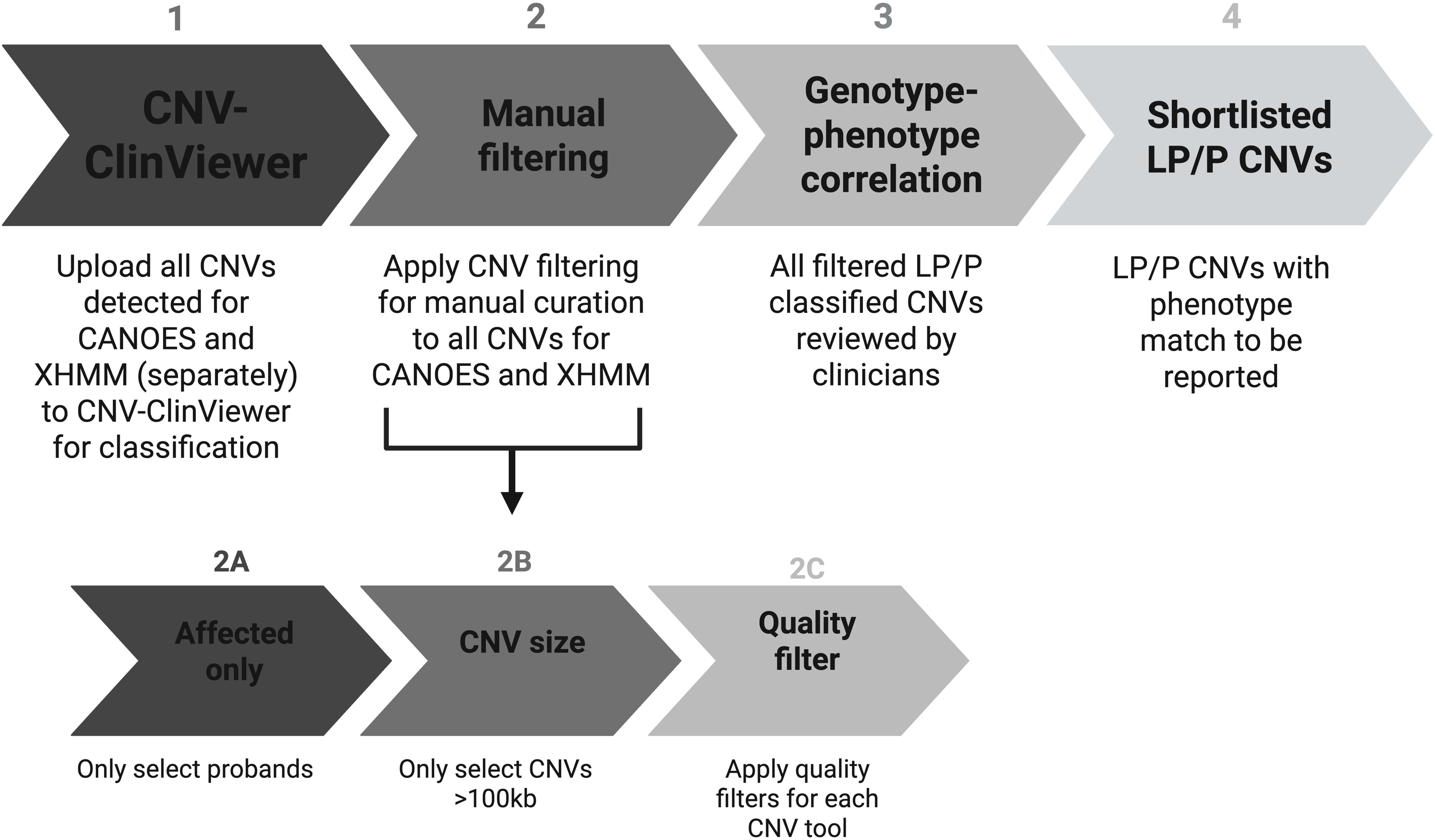
A) The overlap of CNVs detected in probands by the two tools employed. B) CNVs detected are further categorized by size and type (deletion or duplication). Likely pathogenic (LP) and pathogenic (P) are indicated. C) The prioritized shortlist of CNVs of which 75% (33/44) were detected by both tools, these exclude the LP/P CNVs shown in figure 4B which were located on chromosome X and excluded from the shortlist due to low quality metrics.

Fourteen of the 56 LP/P CNV (Figures 4A and 4B) were not included in the final shortlist, these CNV were all detected on the X-chromosome in six different participants with lower quality metrics than seen in the other shortlisted CNV. This could indicate that these were false positive calls and were thus excluded from further analysis. Two CNV smaller than 100kb were included in the shortlist as they were in regions of interest and matched the probands’ phenotypes.

Finally, 44 LP/P CNV were shortlisted from 42 participants of which 31 were deletions and 13 duplications (Table 1). These CNV all met the quality criteria set for the individual tools and 33/44 were detected by both tools (Figure 4C). Two participants had two different CNV in the same region (ID 454302 and ID 491093) suggesting the presence of a single underlying structural rearrangement. Phenotypic details of all participants are listed in Supplementary Table 1. Available ES data from both parents allowed the confirmation of *de novo* inheritance in 27 probands and maternal inheritance for one proband. For one male proband, a CNV on chromosome X, absent from the mother, was assumed to be *de novo.* For the remainder of the variants (N=16) only one parent was available, who did not carry the CNV (Table 1).

**Table 1:**
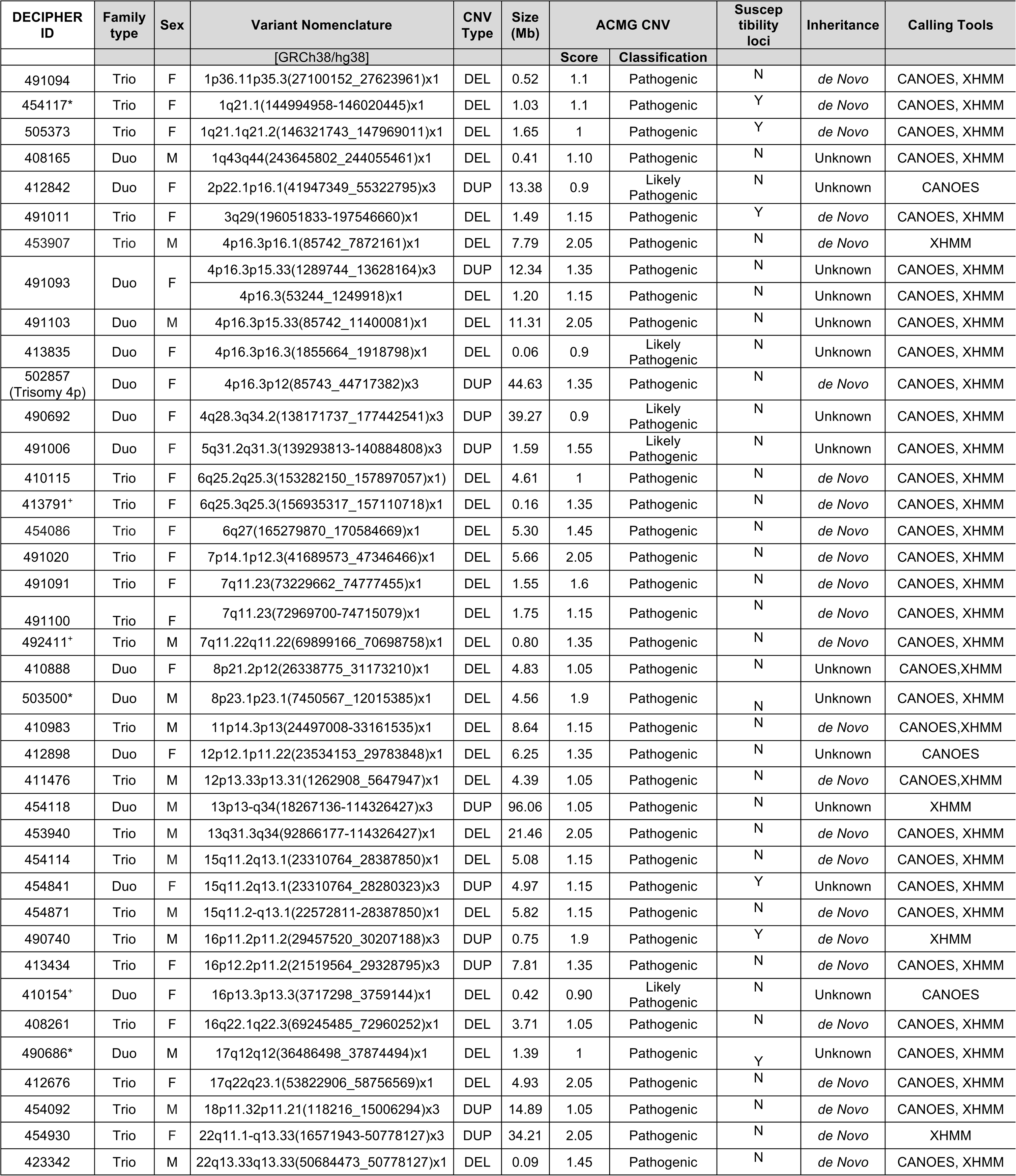
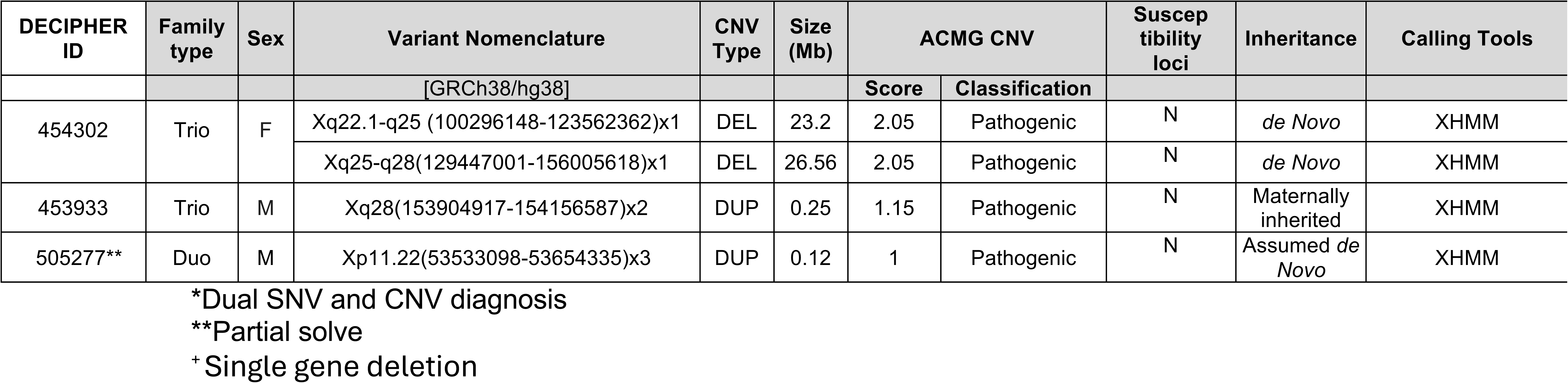
Final shortlist of DDD-Africa CNV detected by CANOES and/or XHMM. Susceptibility loci were defined according to Goh et al., 2025.

Nineteen selected ES-CNV were studied by CMA and all were confirmed. Excluding aneuploidies and more complex rearrangements, the average difference in CNV size between ES and CMA was 309kb (Supplementary Table 2). The differences in size ranged from 10kb to over 1Mb, with CMA reporting a larger CNV size in 15/19 cases. In 12/19 (63%) of the cases, the disease-associated genes identified within the CNV regions were identical between ES and CMA. In the remaining 7/19 (37%) cases, additional disease-associated genes were included in the CNV, but this did not translate into a change in the classification of the CNV or in management for the participant. CMA provided additional clarification for three CNV due to higher resolution and the ability to interpret the dosage more effectively. These included a mosaic trisomy detected on chromosome 22 (ID 454930) and a duplication on chromosome 13 which was identified as a trisomy 13 (ID 454118) as the entire chromosome 13 region was duplicated. Array also confirmed a complex rearrangement (ID 491093) which was detected as separate deletion and duplication events on chromosome 4 after ES CNV analysis.

In this study, three participants harbored a deletion affecting only a single gene. One of these is linked to Rubenstein Taybi syndrome (*CREBBP*) (participant ID 410154), the second to Coffin Siris syndrome (*ARID1B*) (participant ID 413791) and the third CNV is located in the *AUTS2* gene (participant ID 492411), linked to AUTS2-related syndromic intellectual disability.

One of the LP/P CNV not included was a maternally inherited recurrent 15q11.2 deletion (BP1-BP2) which has recently been reported with a very low penetrance, and did not fully explain the phenotype of the participant (31). We also observed well described susceptibility loci in 6/505 probands (incidence 1/84, 1.2%), including the 1q21 deletion (n = 2), 3q29 deletion, 15q11-q13 duplication, 16p11.2 duplication and a 17q12 deletion which were detected as part of the 44 LP/P CNV (Table 1).

## Discussion

In this study, we estimated the value of using CNV detection tools for exome data, in a cohort of 505 probands with unexplained DD from two African countries, South Africa and DRC. We were able to identify disease-causing CNV in 42 probands, producing a diagnostic yield of 8.3%. Other studies have reported an ES-CNV diagnostic yield between 2.3%-20.8% (10, 32–34), with trio-ES studies producing the highest yields. Our findings align with the diagnostic yields reported in these populations, but for a cohort that has limited access to genetic services.

We demonstrated that access to genetics testing influences the diagnostic yield of an ES diagnostic approach. The yield was significantly different (P<0.05) in the two sites, [South Africa with 22/358 (6.1%) and the DRC with 20/147 (13.6%)] which is likely attributable to differences in prior access to diagnostic genetic testing. For South Africa, this is likely indicating the lower boundary of diagnostic yield, which would be higher if CNV analysis were done first line (35).

Compared to CMA with an estimated first-line diagnostic yield of 15-20% (36, 37), and given the limited testing options available in most LMICs, ES presents a potentially cost-effective single test approach to detect both SNVs and CNV. A recent study (38) also supports ES as a cost-effective option for first-tier testing specifically for patients with neurodevelopmental disorders and congenital malformations. Although this study indicated that CNV size estimates were slightly more accurate from CMA data, these differences did not materially affect variant interpretation or classification in the ES dataset. Although ES does not detect all CNV across the genome, it could limit the number of patients requiring CMA if done as a first-tier test. There is thus strong evidence that deploying ES as a first-tier test for patients with unexplained DD could be the most resource efficient approach, an important consideration for resource-constrained environments.

This study demonstrated that employing a combination of CNV detection tools to analyze ES, yields more accurate and reliable results than implementing a single approach. This was evident as neither of the two tools used on the DDD-Africa dataset detected all pathogenic or likely pathogenic CNV individually (Table 1). Some of the variance can be attributed to the tools using different variant detection approaches, for instance XHMM being calibrated to call CNV on sex chromosomes. A total of 68 CNV were detected on the X- and Y chromosomes and four of these CNV were deemed diagnostically relevant (Table 1). Previous studies have also found that combining ES CNV tools could yield superior results (10, 39, 40). Algorithms will undergo further development, resulting in improved detection and accuracy, which may enable the implementation of a single CNV detection tool using NGS data. Until then, it is recommended to implement an ensemble of CNV tools as it would result in improved sensitivity and accuracy.

Most pathogenic CNV detected in this study were larger than 1Mb (Figure 4B) with very few below 500kb which is consistent with findings from previous studies (5, 29, 41). Clinically, CNV are typically classified as pathogenic or likely pathogenic based on several factors, including size thresholds, gene content, *de novo* status, and occurrence in disease cohorts (26). Larger CNV are more likely to disrupt multiple genes or regulatory regions, increasing their potential to cause disease. When implementing CNV calling from ES data, it could be beneficial to start by prioritizing larger CNV (>100kb) and those overlapping known DD associated genes, to focus effort on CNV most easily interpreted as disease-causing.

We observed one maternally inherited LP/P variant in this study. In all cases where exome data from both parents were available (N=27), CNV were confirmed to be *de novo*.

A recent study reported that the African population shows lower odds of carrying specific recurrent CNV or disease-causing CNV (42). This trend was not evident in our study as a number of these were observed in our relatively small sample. The incidence of individual recurrent CNV in previous studies ranged from 0.04% - 0.2% among individuals with DD with a combined incidence for the five susceptibility loci (indicated in Table 1) of ∼0.5% (31, 43). Susceptibility loci have a reduced penetrance, and do not always lead to a disease phenotype. In contrast to high penetrance CNV, susceptibility CNV can be inherited from unaffected or mildly affected parents. In our cohort, in all four informative cases, the susceptibility CNV were *de novo*. A dual diagnosis was observed in two of the six cases (Table 1) [ID 454117 (*MYH3* variant) and ID 490686 (*PPP2R1A* variant)]. The phenotype of these two individuals can be sufficiently explained by the pathogenic SNV involved. One other participant (ID 503500) had a high penetrant CNV (8p23.1 recurrent deletion) and a likely pathogenic SNV detected in the *NF1* gene, consistent with a dual diagnosis.

Interestingly, although 62% (315/505) of index cases in this cohort were male, the majority of participants with a pathogenic or likely pathogenic CNV (59.5%, N=25) identified were female as shown in Table 1. This sex difference was statistically significant in our cohort (p<1×10^−3^). A similar trend has been reported in previous studies showing a higher burden of pathogenic CNV in females compared to males (44–46).

Our study makes an important contribution by providing African data, effectively expanding the knowledge base in public data repositories that is currently predominantly European-focused. This emphasizes a more general issue that there is still a lack of high-quality genomic data especially from Africa which diminishes optimal implementation of genomic medicine efforts and adds to healthcare inequalities (47, 48). This bias complicates comparison across global populations, effective data analysis and ultimately variant classification. More diverse data will greatly improve reclassification of CNV, positively impacting clinical diagnostics globally.

As with all technologies, there are specific limitations to ES CNV detection. Primarily the focus on protein coding exonic regions means not all CNV can be accurately identified with this method. Exact breakpoints and dosage of CNV, small CNV (<1 exon), inversions and translocations as well as mosaicism are not identified as part of this analysis. Additionally, due to the challenges associated with aligning reads in repetitive or structurally complex regions, short-read ES may be prone to false positives or may fail to detect CNV in these genomic contexts. Therefore, long-read sequencing studies incorporating both SNV and CNV analysis, might be preferred in future diagnostic routine testing, further optimizing diagnostic yield (49, 50). Given the substantially higher costs of implementation and analysis associated with long-read sequencing, incorporating ES with CNV analysis as first-tier test for DD is currently still the more cost-effective approach, particularly in LMICs.

## Conclusion

Our study supports the implementation of ES as a first-tier testing strategy in patients with undiagnosed DD, particularly when incorporating CNV analysis. In resource-limited settings, where access to multiple testing strategies such as CMA and karyotyping is often limited, exome-based CNV detection presents a scalable, efficient, and cost-effective first line diagnostic alternative. This study shows that CMA may not be required except to confirm larger, more complex rearrangements presenting as multiple CNV on ES CNV detection methods. It also highlights the significant added value of integrating CNV analysis into routine ES workflows for the diagnosis of DD, particularly in LMICs. Improving the accuracy, reliability, and usability of CNV calling tools will directly enhance diagnostic yield and clinical outcomes, making it a critical focus for future research. At present, combining multiple CNV tools still seems to be more accurate than employing a single bioinformatic tool. Priorities for algorithm development should include increasing sensitivity for clinically relevant CNV, reducing false positives, and ensuring compatibility with diverse sequencing platforms and data quality. Ultimately, embedding robust exome-based CNV detection into standard diagnostic pipelines is not only scientifically and economically justified—it is a necessary and equitable step toward improving genetic care and reducing diagnostic disparities for individuals with DD in LMICs.

## Data availability statement

This research generated genomic data (CNV) and clinical data. The CNV will be submitted to ClinVar. In accordance with funder agreements, the complete dataset from the DDD-Africa study, including all phenotypic data, is available on the European Genome-phenome Archive (EGAS00001008319) to ensure public access.

## Acknowledgements

We gratefully acknowledge the participants of the DDD-Africa study. The authors would like to acknowledge the clinical and genetics team at the Division of Human Genetics, National Health Laboratory Service, and School of Pathology, University of the Witwatersrand, Johannesburg as well the human genetics team at Inkosi Albert Luthuli Central Hospital (IALCH), University of Kwazulu-Natal, Durban and the human genetics team at the Faculty of Medicine of the University of Kinshasa.

## Author Contributions

NL: conceptualization, visualization, data acquisition, data curation and writing–original draft. PM: conceptualization, data acquisition, data curation, writing–review and editing. PTM: data analysis, writing–review and editing. EH, TN and LY: data acquisition, writing–review and editing. GM: data acquisition, writing–review and editing. KVDB: data curation, manuscript review. HVF: data acquisition, conceptualization, manuscript review. ME.H: data acquisition, conceptualization, manuscript review. PLT: conceptualization, funding acquisition, manuscript review. KD: conceptualization, visualization, writing–review and editing. AK: data acquisition, conceptualization, funding acquisition, writing–review and editing. NC: conceptualization, funding acquisition, supervision and writing–review and editing. AL: data acquisition, conceptualization, funding acquisition, writing–review and editing. ZL: conceptualization, funding acquisition, supervision and writing–review and editing.

## Ethical Approval

DDD-Africa received approval from the Human Research Ethics Committee (Medical) of the University of the Witwatersrand (certificate number: M230567) as the main site and from the the University of Kinshasa (reference number: ESP/CE/050/2018). Informed consent was obtained from all participants as per IRB regulations and all individual level data was de-identified by the use of a code and numbering system.

## Supplementary material

Supplementary Table 1

Supplementary Table 2

