## Supplemental Table 2 for "Copy Number Variant analysis by exome sequencing is an effective approach to optimize diagnostic yield for developmental disorders – the DDD-Africa study"

**Supplementary Table 2: Comparison of CNV coordinates and sizes from ES data and CMA.**

| Patient ID | Exome CNV (GRCh38) | Size (kb) | Array CNV | Size (kb) | Size Difference (kb) |
|---|---|---|---|---|---|
| 413835 | 4:1855664-1918798 | 63 | 4:1875256-1925567 | 50 | 10 |
| 410888 | 8:26338775-31173210 | 4834 | 8:26317222-31242903 | 4926 | 92 |
| 453940 | 13:92866177-114326427 | 21460 | 13:93105502-115093155 | 21988 | 527 |
| 454092 | 18:118216-15006294 | 14888 | 18:12989-15325198 | 15312 | 424 |
| 454114 | 15:23310764_28280323 | 4970 | 15:23300226-28780204 | 5480 | 510 |
| 454871 | 15:22572811-28387850 | 5815 | 15:22684152-28846172 | 6162 | 347 |
| 491011 | 3:196051833-197546660 | 1495 | 3:195740402-197334627 | 1594 | 99 |
| 408261 | 16:69245485-72960252 | 3715 | 16:69254089-73997817 | 4744 | 1029 |
| 410115 | 6:153282150-157897057 | 4615 | 6:153598844-158301597 | 4703 | 88 |
| 410983 | 11:24497008-33161535 | 8665 | 11:24381118-33162974 | 8782 | 117 |
| 411476 | 12:1262908-5647947 | 4385 | 12:1388453-5685697 | 4297 | 88 |
| 412676 | 17:53822906-58756569 | 4934 | 17:51714213-56978556 | 5264 | 331 |
| 412842 | 2:41947349-55322795 | 13375 | 2:41911045-55276923 | 13366 | 10 |
| 412898 | 12:23534153-29783848 | 6250 | 12:22882439-30246209 | 7364 | 1114 |
| 423342 | 22:50684473-50778127 | 94 | 22:50685063-50739836 | 55 | 39 |
| 505277 | X:53533098-53654335 | 121 | X:53532771-53764209 | 231 | 110 |
| 454302 | X:100296148-156005618 | 55709 | X:100296148-123562362 and X:129447001-156005618 | | |
| 454118 | 13:18267136-114326427 | 96059 | trisomy 13 | | |
| 454930 | 22:16571943-50778127 | 34206 | arr(22)x2~3 Mosaic trisomy | | |
