## Supplemental Table 1 for "Copy Number Variant analysis by exome sequencing is an effective approach to optimize diagnostic yield for developmental disorders – the DDD-Africa study"

Supplementary Table 1: Phenotypes of the 42 CNV positive patients presented as Human Phenotype Ontology (HPO) terms.

| **Patient ID** | **Clinical term** |
| --- | --- |
| 408165 | HP:0011344 Severe global developmental delay  HP:0000750 Delayed speech and language development HP:0000252 Microcephaly  HP:0001348 Brisk reflexes  HP:0000347 Micrognathia HP:0000028 Cryptorchidism HP:0001537 Umbilical hernia  HP:0001999 Dysmorphic facial features |
| 408261 | HP:0011343 Moderate global developmental delay HP:0000750 Delayed speech and language development HP:0001643 Patent ductus arteriosus HP0000486 Strabismus HP:0000767 Pectus excavatum HP:0002650 Scoliosis HP:0001382 Joint hypermobility  HP:0001999 Dysmorphic facial features |
| 410115 | HP:0011344 Severe global developmental delay HP:0000750 Delayed speech and language development  HP:0000280 Coarse facial features HP:0001537 Umbilical hernia  HP:0001999 Dysmorphic facial features |
| 410154 | HP:0011343 Moderate global developmental delay HP:0000750 Delayed speech and language development HP:0001643 Patent ductus arteriosus HP:0001488 Bilateral ptosis HP:0000347 Micrognathia HP:0000218 High palate  HP:0001156 Brachydactyly  HP:0001999 Dysmorphic facial features |
| 410888 | HP:0011343 Moderate global developmental delay HP:0000750 Delayed speech and language development HP:0000252 Microcephaly  HP:0002283 Global brain atrophy HP:0001999 Dysmorphic facial features |
| 410983 | HP:0011344 Severe global developmental delay HP:0000750 Delayed speech and language development  HP:0000526 Aniridia  HP:0025502 Overweight  HP:00005189 Cataract  HP:0000028 Cryptorchism  HP:0001999 Dysmorphic facial features |
| 411476 | HP:0000750 Delayed speech and language development HP:0011344 Severe global developmental delay  HP:0000252 Microcephaly  HP:0001999 Dysmorphic facial features |
| 412676 | HP:0011344 Severe global developmental delay HP:0001344 Absent speech HP:0000252 Microcephaly HP:0001999 Dysmorphic facial features |
| 412842 | HP:0011344 Severe global developmental delay HP:0001344 Absent speech  HP:0000280 Coarse facial features HP:0002059 Cerebral atrophy HP0001348 Brisk reflexes HP0100704 Cortical visual impairment HP:0001537 Umbilical hernia  HP:0001999 Dysmorphic facial features |
| 412898 | HP:0011343 Moderate global developmental delay HP:0000750 Delayed speech and language development HP:0000252 Microcephaly  HP:0001508 Failure to thrive HP:0001348 Brisk reflexes  HP:0001999 Dysmorphic facial features |
| 413434 | HP:0011343 Moderate global developmental delay HP:0000750 Delayed speech and language development HP:0003691 Scapular winging HP:0003307 Hyperlordosis HP:0002987 Elbow flexion contracture  HP:0001999 Dysmorphic facial features |
| 413791 | HP:0011343 Moderate global developmental delay HP:0000750 Delayed speech and language development HP:0001252 Muscular hypotonia HP:0001250 Seizures HP:0000545 Myopia HP:0000486 Strabismus HP:0001537 Umbilical hernia  HP:0002719 Recurrent infections  HP:0001999 Dysmorphic facial features |
| 413835 | HP:0011342 Mild global developmental delay HP:0001537 Umbilical hernia  HP:0001999 Dysmorphic facial features |
| 423342 | HP:0000750 Delayed speech and language development HP:0011344 Severe global developmental delay HP:0001212 Prominent fingertip pads  HP:0001999 Dysmorphic facial features |
| 453907 | HP:0002194 Delayed gross motor development HP:0000750 Delayed speech and language development HP:0010864 Intellectual disability, severe  HP:0010714 2-4 toe syndactyly HP:0001776 Bilateral talipes equinovarus HP:0001250 Seizures HP:0012171 Stereotypical hand wringing HP:0000154 Wide mouth  HP:0001999 Dysmorphic facial features |
| 453933 | HP:0000750 Delayed speech and language development  HP:0002342 Intellectual disability, moderate HP:0001034 Hypermelanotic macule HP:0006610 Wide intermamillary distance  HP:0000325 Triangular face  HP:0001999 Dysmorphic facial features |
| 453940 | HP:0002194 Delayed gross motor development HP:0000750 Delayed speech and language development  HP:0006143 Abnormal finger flexion crease HP:0007598 Bilateral single transverse palmar creases HP:0000028 Cryptorchidism HP:0000400 Macrotia HP:0000742 Self-mutilation HP:0012171 Stereotypical hand wringing |
| 453940 | HP:0010804 Tented upper lip vermilion HP:0006610 Wide intermamillary distance  HP:0000470 Short neck  HP:0011081 Incisor macrodontia  HP:0001999 Dysmorphic facial features |
| 454086 | HP:0010862 Delayed fine motor development HP:0000750 Delayed speech and language development  HP:0010864 Intellectual disability, severe  HP:0000252 Microcephaly  HP:0000020 Urinary incontinence  HP:0000752 Hyperactivity  HP:0020045 Esodeviation  HP:0410030 Cleft lip HP:0000324 Facial asymmetry HP:0001999 Dysmorphic facial features |
| 454092 | HP:0010862 Delayed fine motor development HP:0002194 Delayed gross motor development HP:0000750 Delayed speech and language development HP:0001256 Intellectual disability, mild  HP:0000252 Microcephaly  HP:0002251 Aganglionic megacolon HP:0009836 Broad distal phalanx of finger HP:0001836 Camptodactyly of toe HP:0012622 Chronic kidney disease HP:0001999 Dysmorphic facial features |
| 454114 | HP:0002194 Delayed gross motor development  HP:0000252 Microcephaly  HP:0000244 Brachyturricephaly HP:0000028 Cryptorchidism HP:0001290 Generalized hypotonia HP:0001513 Obesity HP:0000954 Single transverse palmar crease HP:0001770 Toe syndactyly  HP:0000414 Bulbous nose  HP:0001999 Dysmorphic facial features |
| 454117 | HP:0008936 Axial hypotonia HP:0011260 Darwin notch of helix HP:0000293 Full cheeks HP:0002162 Low posterior hairline HP:0007413 Nevus flammeus of the forehead HP:0005989 Redundant neck skin HP:0000331 Short chin HP:0000470 Short neck HP:0001762 Talipes equinovarus HP:0009909 Uplifted earlobe HP:0006610 Wide intermamillary distance HP:0040149 Woolly scalp hair  HP:0001999 Dysmorphic facial features |
| 454118 | HP:0001290 Generalized hypotonia HP:0002240 Hepatomegaly HP:0009890 High anterior hairline HP:0007413 Nevus flammeus of the forehead HP:0001539 Omphalocele HP:0011682 Perimembranous ventricular septal defect HP:0001162 Postaxial hand polydactyly HP:0002092 Pulmonary arterial hypertension HP:0012745 Short palpebral fissure HP:0001762 Talipes equinovarus HP:0001770 Toe syndactyly HP:0006610 Wide intermamillary distance  HP:0001999 Dysmorphic facial features |
| 454302 | HP:0010862 Delayed fine motor development  HP:0000252 Microcephaly  HP:0002056 Abnormality of the glabella HP:0100490 Camptodactyly of finger HP:0410030 Cleft lip HP:0040018 Clinodactyly of hallux HP:0004681 Deep longitudinal plantar crease HP:0002194 Delayed gross motor development HP:0000750 Delayed speech and language development HP:0000218 High palate HP:0000767 Pectus excavatum HP:0001763 Pes planus HP:0012428 Prominent calcaneus HP:0000470 Short neck HP:0005659 Thoracic kyphoscoliosis HP:0001770 Toe syndactyly HP:0006610 Wide intermamillary distance  HP:0001999 Dysmorphic facial features |
| 454841 | HP:0002342 Intellectual disability, moderate  HP:0000752 Hyperactivity HP:0000664 Synophrys HP:0000325 Triangular face HP:0000582 Upslanted palpebral fissure  HP:0001999 Dysmorphic facial features |
| 454871 | HP:0002194 Delayed gross motor development HP:0000750 Delayed speech and language development HP:0010864 Intellectual disability, severe  HP:0020049 Exodeviation HP:0000752 Hyperactivity HP:0001250 Seizures  HP:0000324 Facial asymmetry  HP:0001999 Dysmorphic facial features |
| 454930 | HP:0000252 Microcephaly  HP:0001679 Abnormal aortic morphology HP:0000062 Ambiguous genitalia HP:0002023 Anal atresia HP:0009892 Anotia HP:0030021 Auricular tag HP:0000337 Broad forehead HP:0410030 Cleft lip HP:0000175 Cleft palate HP:0000086 Ectopic kidney HP:0002816 Genu recurvatum HP:0001034 Hypermelanotic macule HP:0000316 Hypertelorism HP:0000612 Iris coloboma HP:0004626 Lumbar scoliosis HP:0008551 Microtia HP:0010733 Naevus flammeus of the eyelid HP:0001319 Neonatal hypotonia HP:0001643 Patent ductus arteriosus HP:0001763 Pes planus HP:0004451 Postauricular skin tag HP:0008678 Renal hypoplasia/aplasia HP:0030799 Scaphocephaly HP:0009381 Short finger HP:0000954 Single transverse palmar crease HP:0001750 Single ventricle HP:0001770 Toe syndactyly HP:0001824 Weight loss HP:0000260 Wide anterior fontanel  HP:0000470 Short neck  HP:0001999 Dysmorphic facial features |
| 490686 | HP:0011344 Severe global developmental delay  HP:0000750 Delayed speech and language development HP:0001999 Dysmorphic facial features |
| 490692 | HP:0012736 Profound global developmental delay  HP:0001344 Absent speech HP:0000252 Microcephaly HP:0001250 Seizures  HP:0001999 Dysmorphic facial features |
| 490740 | HP:0011342 Mild global developmental delay  HP:0000750 Delayed speech and language development HP:0008936 Muscular hypotonia of the trunk  HP:0000028 Cryptorchidism  HP:0001999 Dysmorphic facial features |
| 491006 | HP:0010862 Delayed fine motor development HP:0002194 Delayed gross motor development  HP:0010864 Intellectual disability, severe  HP:0011918 Clinodactyly of the 4th toe HP:0001864 Clinodactyly of the 5th toe HP:0011261 Darwin tubercle of helix HP:0000699 Diastema HP:0004373 Focal dystonia HP:0000752 Hyperactivity HP:0011081 Incisor macrodontia HP:0100807 Long fingers HP:0001847 Long hallux HP:0000322 Short philtrum HP:0012471 Thick vermilion border  HP:0000276 Long face  HP:0001999 Dysmorphic facial features |
| 491011 | HP:0010862 Delayed fine motor development HP:0000750 Delayed speech and language development  HP:0002342 Intellectual disability, moderate HP:0010529 Echolalia HP:0100962 Excessive shyness HP:0000752 Hyperactivity HP:0008551 Microtia  HP:0001999 Dysmorphic facial features |
| 491020 | HP:0010862 Delayed fine motor development HP:0002194 Delayed gross motor development HP:0000750 Delayed speech and language development  HP:0010712 1-4 toe syndactyly HP:0032190 Abnormal meniscus morphology HP:0001018 Abnormal palmar dermatoglyphics HP:0001274 Agenesis of corpus callosum HP:0002506 Diffuse cerebral atrophy HP:0020045 Esodeviation HP:0002553 Highly arched eyebrow HP:0000238 Hydrocephalus HP:0011081 Incisor macrodontia HP:0010864 Intellectual disability, severe HP:0002751 Kyphoscoliosis HP:0001520 Large for gestational age HP:0100807 Long fingers HP:0001845 Overlapping toe HP:0001841 Preaxial foot polydactyly HP:0004482 Relative macrocephaly HP:0005676 Rudimentary postaxial polydactyly of hands HP:0002652 Skeletal dysplasia HP:0001744 Splenomegaly HP:0012171 Stereotypical hand wringing HP:0012471 Thick vermilion border HP:0001537 Umbilical hernia HP:0006610 Wide intermamillary distance  HP:0002000 Short columella HP:0000470 Short neck HP:0001999 Dysmorphic facial features |
| 491091 | HP:0002194 Delayed gross motor development HP:0000750 Delayed speech and language development  HP:0002342 Intellectual disability, moderate HP:0000718 Aggressive behavior  HP:0000805 Enuresis HP:0000752 Hyperactivity HP:0100716 Self-injurious behaviour  HP:0001999 Dysmorphic facial features |
| 491093 | HP:0002194 Delayed gross motor development HP:0000750 Delayed speech and language development HP:0000752 Hyperactivity HP:0001256 Intellectual disability, mild HP:0000400 Macrotia HP:0001763 Pes planus HP:0100716 Self-injurious behaviour HP:0000736 Short attention span  HP:0001999 Dysmorphic facial features |
| 491094 | HP:0002194 Delayed gross motor development HP:0000750 Delayed speech and language development  HP:0010864 Intellectual disability, severe  HP:0002066 Gait ataxia HP:0000752 Hyperactivity HP:0008551 Microtia HP:0000508 Ptosis HP:0006610 Wide intermamillary distance  HP:0000324 Facial asymmetry  HP:0001999 Dysmorphic facial features |
| 491100 | HP:0002194 Delayed gross motor development HP:0000750 Delayed speech and language development  HP:0000256 Macrocephaly  HP:0000718 Aggressive behavior HP:0025112 Auditory sensitivity HP:0000752 Hyperactivity HP:0000400 Macrotia HP:0030799 Scaphocephaly HP:0000736 Short attention span HP:0012171 Stereotypical hand wringing  HP:0001999 Dysmorphic facial features |
| 491103 | HP:0002194 Delayed gross motor development  HP:0000252 Microcephaly  HP:0008936 Axial hypotonia HP:0001156 Brachydactyly HP:0000518 Cataract HP:0006191 Deep palmar crease HP:0005469 Flat occiput HP:0000612 Iris coloboma HP:0000400 Macrotia HP:0000520 Proptosis HP:0000278 Retrognathia HP:0000470 Short neck HP:0002209 Sparse scalp hair |
| 492411 | HP:0011343 Moderate global developmental delay  HP:0000252 Microcephaly  HP:0008936 Muscular hypotonia of the trunk  HP:0003502 Mild short stature  HP:0001999 Dysmorphic facial features |
| 502857 | HP:0011343 Moderate global developmental delay HP:0001344 Absent speech HP:0000252 Microcephaly HP:0004322 Short stature  HP:001252 Muscular hypotonia  HP:0002781 Upper airway obstruction  HP:0001999 Dysmorphic facial features |
| 503500 | HP:0000750 Delayed speech and language development HP:0011343 Moderate global developmental delay.  HP:0001537 Umbilical hernia  HP:0030052 Inguinal freckling  HP:000997 Axillary freckling  HP:0000957 Cafe-au-lait spot  HP:0001999 Dysmorphic facial features |
| 505277 | HP:0011342 Mild global developmental delay  HP:0000750 Delayed speech and language development HP:0004381 Supravalvular aortic stenosis  HP:0001999 Dysmorphic facial features |
| 505373 | HP:0000252 Microcephaly HP:0011679 Tetralogy of Fallot with pulmonary stenosis  HP:0004325 Decreased body weight HP:0001508 Failure to thrive HP:010025 Postaxial polydactyly  HP:0001999 Dysmorphic facial features |
